# Skin diseases, including skin NTDs, identified through community-based integrated skin screening: Findings from the PEP4LEP study in Tanzania

**DOI:** 10.64898/2025.12.02.25341087

**Authors:** N. Mwageni, A. Schoenmakers, R. van Wijk, D.V. Kamara, R. M. Kisonga, B. Njako, P. Nyakato, L. Mieras, S. J. de Vlas, J. H. Richardus, C. Kasang, E. J. Masenga, S. E. Mshana

**Author notes:** Corresponding author: Anne Schoenmakers. Contributed equally as first author (N. Mwageni, A. Schoenmakers).

## Abstract

**Background:** Skin diseases, including skin-related neglected tropical diseases (NTDs), contribute substantially to the global disease burden, particularly in low- and middle-income countries. Strengthening community health programmes to promote early diagnosis and treatment of skin conditions is essential. This study describes skin diseases identified during community-based integrated skin screening events conducted as part of the PEP4LEP project in three districts in Tanzania. A second objective is to use prevalence data to inform integrated skin health and NTD strategies within Tanzania and in similar contexts beyond.

**Methodology:** A cross-sectional study was conducted within the PEP4LEP implementation research project between December 2019 and March 2024. Community ‘skin camps’ were organised near leprosy index patients – without disclosing their disease status – to screen contacts from the 20 nearest households and administer preventive medication to those eligible. A dermatologist screened participants and provided medical advice, medication (prescriptions) and/or referrals. Demographic data and diagnoses were registered. A subset of participants also completed an additional questionnaire. Paper-based data were entered into REDCap and Microsoft Excel, and analysed in SPSS and Stata using descriptive statistics and Chi-square tests.

**Results:** Of the 4,501 persons screened during 44 community skin camps, 2,527 (56.1%) were diagnosed with one or more skin diseases, of whom 392 (15.5%) had multiple conditions. Among 2,959 recorded diagnoses, 92 distinct skin diseases were identified. The most common conditions were tinea capitis (24.0% of diagnoses), pityriasis versicolor (13.0%), and scabies (7.3%). Five skin NTDs were seen among 280 participants, accounting for 6.2% of all screened individuals and 9.5% of all diagnoses: scabies, leprosy (1.5% of diagnoses), onchocerciasis (0.5%), lymphatic filariasis (0.1%), and chromoblastomycosis (0.03%). Of the 1,841 dermatological patients who completed the additional questionnaire, 28.3% had previously sought medical care, with over half (54.6%) consulting drug-selling shops.

**Conclusion:** Over half of the community members screened in Tanzanian skin camps had at least one skin condition, with skin NTDs accounting for almost 10% of diagnoses. Strengthening integrated, community-based dermatological and NTD control approaches is vital to improve early detection and treatment, particularly in underserved areas.

## Introduction

Skin conditions are among the most common health problems globally, affecting an estimated 1.8 billion people at any given time, placing a heavy burden on individuals and healthcare systems [1–6]. The burden is especially high in low- and middle-income countries [7]. Factors such as warm and humid environments, limited access to care, overcrowding, and poor water, sanitation and hygiene (WASH) infrastructure exacerbate vulnerability [5,7–9]. While associated mortality is low, many skin diseases can cause significant morbidity, stigma, and psychosocial distress – particularly neglected tropical diseases (NTDs) that manifest on the skin, such as leprosy, lymphatic filariasis, and onchocerciasis [3,10–13]. According to the World Health Organization (WHO) estimates, skin NTDs account for up to 10% of the dermatological burden in tropical and resource-limited settings [4]. Many of these diseases can lead to disability if not detected and treated early, making timely diagnosis and intervention critical [2,3,10–12,14]. Despite this burden, research efforts and funding for skin diseases remain disproportionately low compared with their impact [3,15].

Dermatologic conditions can frequently be diagnosed by visual inspection [14,16]. Skin examination, a low-cost and non-invasive intervention, offers a platform for early detection particularly in settings where other specialised health services are scarce. Since half of the NTDs manifest on the skin, integrated skin examination offers an efficient way to identify multiple conditions in a single assessment [4,10,17,18]. The integrated approach is endorsed by the WHO’s strategic framework for integrated control and management of skin NTDs and in their NTD road map towards 2030 [19,20].

Currently, most dermatological literature, educational materials, and digital diagnostic tools predominantly focus on light skin tones, with limited attention to the distinct clinical features and varying presentations of skin conditions in skin of colour [21–25]. At the same time, several dermatoses, such as pseudofolliculitis barbae, papulosa nigra and keloid, occur more frequently or almost exclusively in individuals with black or brown skin linked to structural, genetic, and immunological variation [9,21,23]. Yet these conditions remain insufficiently represented in global research and clinical resources, ultimately contributing to inequitable care [9,14,21–23].

Another gap in the existing literature is that most studies on skin disease prevalence in African countries have been facility-based or limited to specific subgroups such as school-aged children [26,27]. In Tanzania, community-based data remain sparse with only two community studies from the 1990s and one survey among Maasai adults attending an outreach clinic in 2019 identified [8,28,29]. Current data on the burden of skin diseases – particularly skin NTDs – across all age groups in rural Tanzanian communities remain limited, hindering the design of targeted and integrated health interventions.

To help address these gaps, the present study was conducted as part of the PEP4LEP project, an implementation study in Ethiopia, Mozambique, and Tanzania that combines community-based skin screening with the provision of leprosy chemoprophylaxis [30]. The project compares two approaches: (1) community-based ‘skin camps’, where community contacts of a leprosy index patient are screened for leprosy and other skin diseases, and given a single dose of rifampicin when eligible, and (2) health centre-based screening, offering the same health intervention to household contacts of enrolled index patients. A modelling study using PEP4LEP skin camp data suggested that such camps can improve early leprosy detection and thereby reduce transmission and future incidence [31]. This paper examines a PEP4LEP cohort from Tanzania’s community-based skin camps, where participants received integrated skin screening by a dermatologist. The specific objectives of this cohort study were to:

1. Determine the prevalence and spectrum of skin diseases – including skin NTDs – identified during community-based integrated skin screening in rural Tanzania;
2. Generate evidence to inform strategies for strengthening integrated skin health and NTD services, and to support advocacy and policymaking for scalable, community-based approaches in similar contexts.

The recently adopted World Health Assembly (WHA) Resolution WHA78.15 recognises skin diseases as a global public health priority and calls for their integration into universal health coverage, improved access to affordable diagnosis and treatment, and strengthened training for frontline health workers [15]. These priorities align closely with the objectives of the current study and the broader aims of the project.

## Methods

### Study design, area and period

A cross-sectional study was conducted between December 2019 and March 2024 in three Tanzanian districts implementing PEP4LEP activities: Morogoro Rural, Mvomero, and Lindi Rural (Figure 1) [30]. These districts are located in south-eastern Tanzania, where most residents depend on agriculture, fishing, and livestock keeping [32,33].

**Figure 1.**
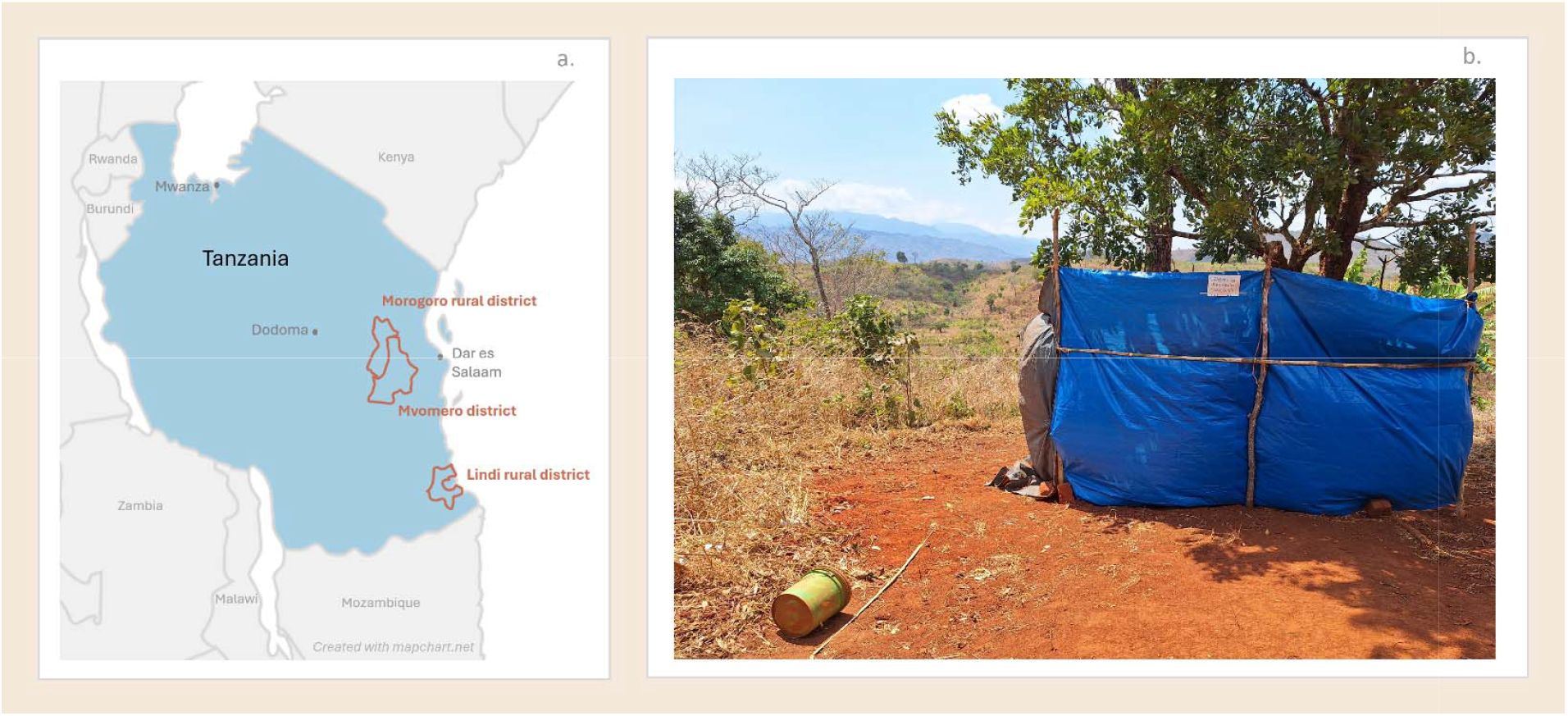
a. Study districts in Tanzania b. Pop-up skin screening station in Tanzanian ‘skin camp’ with a sign reading: “schemu ya uchunguzi (daktari)”, translated: “examination section (doctor)”. Photo ©AS.

### Study population

The study population comprised contacts of leprosy patients who had lived for at least three months in the 20 closest households surrounding the home of a recently diagnosed index patient (diagnosed within six months before inclusion). After obtaining consent from index patients and approval from local authorities and community leaders, community skin screening events (‘skin camps’) were organized [30]. To prevent stigma, the identity of the index patient was not disclosed [30,34]. Community contacts were invited to participate and screened for skin conditions, including skin NTDs. Those eligible were also offered leprosy preventive medication, as the area is endemic for leprosy. The purpose of the intervention was explained to participants through community sensitisation held before the screening events and again at the time of screening. Eligible participants were enrolled after providing informed consent. Contacts were excluded if they, or their legal guardians could not comprehend the purpose and implications of participating in the PEP4LEP study. A more detailed description of the PEP4LEP project can be found in the protocol article [30].

### Skin screening and procedures

In PEP4LEP, dermatologists or trained health workers perform skin screening during community skin camps [30]. For this study, only events with a dermatologist present were included. Screening followed WHO standards to ensure privacy and confidentiality [30,35,36]. Examinations were conducted in well-lit indoor or covered outdoor spaces (Figure 1). Participants were asked to undress for skin inspection. The dermatologist leading the screenings (NM) has 15 years of medical experience, including seven years as dermatologist.

Following the examination, community contacts with skin diseases received medical advice, topical treatment, prescriptions, or medical referrals as appropriate. A single dose of rifampicin (SDR) was provided as post-exposure prophylaxis (PEP) for leprosy to eligible contacts, according to age and weight, following WHO guidance [30,35,36]. Contacts who consented but were not eligible for SDR-PEP were also offered skin screening [30].

### Data collection

Basic demographic information, including sex and age, was recorded on a paper-based data collection form for all individuals screened. For those diagnosed with skin diseases, the specific diagnosis was also documented. For a subset of participants – those enrolled up to July 2023 – an additional questionnaire was administered to collect further details on their condition, symptoms experienced, affected family members and health-seeking behaviour.

### Data processing and analysis

The main participants’ data were entered into REDCap [37], a secure web-based data management system, while information from the additional questionnaire was recorded in Microsoft Excel. Data were analysed using SPSS v26 and Stata v15.1. Continuous variables were summarised using measures of central tendency, and categorical variables were presented as frequencies and percentages. Statistical testing was conducted using the Chi-square test with a 5% significance level.

### Ethical considerations

Permission to conduct the study was approved by the joint CUHAS/BMC Research Ethics and Review Committee (clearance no. CREC/364/2019) [30]. The PEP4LEP project in Tanzania was approved by the National Institute for Medical Research (NIMR) of Tanzania (reference number; NIMR/HQ/R.8c/Vol.1/1530). Consent was signed by the participants or (co-signed) by parents or legal guardians if contacts were below 18 years old. Confidentiality was ensured during the screening of skin conditions by providing a private space. During data processing, personal information was pseudonymised by using coding instead of names. If follow-up care was needed, referrals or prescriptions were provided. PEP4LEP (2.0) is registered in the Pan African Clinical Trials Registry (PACTR), registration number PACTR202303742093429 (original PEP4LEP project registration: Netherlands Trial Register [NTR] registration number NL7294, NTR7503) [38].

### Patient and public involvement

Skin camps were organised in close collaboration with the Ministry of Health, local policy makers, community leaders and community healthcare workers to ensure engagement and acceptability. In addition, a person affected by leprosy is represented in the projects’ International Scientific Steering Committee, ensuring patient perspectives are integrated throughout the project.

## Results

### Characteristics of the participants

A total of 4,501 participants were screened for skin conditions in 44 skin health events in the three districts that implemented PEP4LEP activities in Tanzania during which a dermatologist was present. On average, 102 contacts per event were screened. The median age of all screened contacts was 20 years (IQR 8-42) and among them, 2,646 (58.8%) were females. The characteristics of the participants are summarized in Table 1.

**Table 1.** Characteristics of participants.

| Category | Total population screened |  | Participants with skin conditions |  |  |
| --- | --- | --- | --- | --- | --- |
|  | Number | % | Number | % of total population screened | % persons with skin conditions |
| **Total** | **4,501** | **100%** | **2,527** | **56.1%** | **100%** |
| **Median age (IQR)** | 20 (8-42) |  | 18 (8-38) |  |  |
| **Mean age (95% CI)** | 27.0 (26.3-27.6) |  | 25.0 (24.2-25.3) |  |  |
| **Age groups** |  |  |  |  |  |
| 0-14 | 1,890 | 42.0 | 1,117 | 24.8 | 44.2 |
| 15-29 | 842 | 18.7 | 517 | 11.5 | 20.5 |
| 30-44 | 715 | 15.9 | 398 | 8.8 | 15.8 |
| 45-59 | 483 | 10.7 | 237 | 5.3 | 9.4 |
| ≥60 | 571 | 12.7 | 258 | 5.7 | 10.2 |
| **Sex** |  |  |  |  |  |
| Females | 2,646 | 58.8 | 1,435 | 31.9 | 56.8 |
| Males | 1,855 | 41.2 | 1,091 | 24.2 | 43.3 |

Abbreviations: IQR = interquartile range, CI = confidence interval

### Skin diseases diagnosed

In total, 2,959 skin conditions were identified among 2,527 participants (56.1% of all contacts screened). Nearly 85% (n=2,135) of these participants had a single skin disease, while 15.6% (n=395) received multiple diagnoses (Table 2).

**Table 2.** Number of skin conditions seen per participant.

| Number (n) of skin conditions seen per participant | Number of participants | % of all participants screened (n=4,501) | % of participants with skin conditions (n=2,527) |
| --- | --- | --- | --- |
| 0 | 1,974 | 43.9 | - |
| 1 | 2,132 | 47.4 | 84.4 |
| 2 | 359 | 8.0 | 14.2 |
| 3 | 35 | 0.8 | 1.4 |
| 4 | 1 | 0.02 | 0.04 |

In total, 92 different skin conditions were registered. The top three most encountered skin diseases were: tinea capitis (n=710; 24.0% of all diagnoses; 28.1% of contacts with skin diseases; 15.8% of all contacts screened), pityriasis versicolor (n=384; 13.0% of all diagnoses; 15.2% of contacts with skin diseases; 8.5% of all contacts screened) and scabies (n=217; 7.3% of all diagnoses; 8.6% of contacts diagnosed with skin diseases; 4.8% of all contacts screened). When categorised, fungal diseases were the most common accounting for 56.6% of diagnoses (Table 3).

**Table 3.** Skin diseases detected.

| # | Diagnosis | n | % of all diagnoses made, n=2,959 | % of people diagnosed with skin conditions, n=2,527 | % of all people screened, n=4,501 | # | Diagnosis | n | % of all diagnoses made, n=2,959 | % of people diagnosed with skin conditions, n=2,527 | % of all people screened, n=4,501 |
| --- | --- | --- | --- | --- | --- | --- | --- | --- | --- | --- | --- |
| 1 | Tinea capitis (scalp ringworm)<sup>a</sup> | 710 | 24.0 | 28.1 | 15.8 | 28 | Keloid scar<sup>f</sup> | 14 | 0.5 | 0.6 | 0.3 |
| 2 | Pityriasis versicolor (tinea versicolor)<sup>a</sup> | 384 | 13.0 | 15.2 | 8.5 | 29 | Ulcer(s)<sup>f</sup> | 14 | 0.5 | 0.6 | 0.3 |
| 3 | Scabies<sup>b*</sup> | 217 | 7.3 | 8.6 | 4.8 | 30 | Pellagra<sup>f</sup> | 11 | 0.4 | 0.4 | 0.2 |
| 4 | Tinea cruris<sup>a</sup> | 213 | 7.2 | 8.4 | 4.7 | 31 | Pruritus – otherwise<sup>f</sup> | 10 | 0.3 | 0.4 | 0.2 |
| 5 | Tinea corporis (body ringworm)<sup>a</sup> | 204 | 6.9 | 8.1 | 4.5 | 32 | Pruritic papular eruption (PPE)<sup>e</sup> | 9 | 0.3 | 0.4 | 0.2 |
| 6 | Acne<sup>e</sup> | 130 | 4.4 | 5.1 | 2.9 | 33 | Keratosis pilaris<sup>f</sup> | 7 | 0.2 | 0.3 | 0.2 |
| 7 | Eczema – n.f.d.<sup>e</sup> | 118 | 4.0 | 4.7 | 2.6 | 34 | Dermatosis papulosa nigra<sup>f</sup> | 6 | 0.2 | 0.2 | 0.1 |
| 8 | Atopic dermatitis (atopic eczema)<sup>e</sup> | 101 | 3.4 | 4.0 | 2.2 | 35 | Lichen planus<sup>e</sup> | 6 | 0.2 | 0.2 | 0.1 |
| 9 | Urticaria (hives)<sup>e</sup> | 84 | 2.8 | 3.3 | 1.9 | 36 | Naevus – n.f.d.<sup>f</sup> | 6 | 0.2 | 0.2 | 0.1 |
| 10 | Tinea pedis (athlete's foot)<sup>a</sup> | 66 | 2.2 | 2.6 | 1.5 | 37 | Cheilitis angularis<sup>a</sup> | 5 | 0.2 | 0.2 | 0.1 |
| 11 | Folliculitis<sup>c</sup> | 58 | 2.0 | 2.3 | 1.3 | 38 | Ichthyosis<sup>f</sup> | 5 | 0.2 | 0.2 | 0.1 |
| 12 | Impetigo<sup>c</sup> | 46 | 1.6 | 1.8 | 1.0 | 39 | Pityriasis alba<sup>e</sup> | 5 | 0.2 | 0.2 | 0.1 |
| 13 | Leprosy<sup>c*</sup> | 45 | 1.5 | 1.8 | 1.0 | 40 | Psoriasis<sup>e</sup> | 5 | 0.2 | 0.2 | 0.1 |
| 14 | Contact dermatitis (contact eczema)<sup>e</sup> | 44 | 1.5 | 1.7 | 1.0 | 41 | Senile dermatitis<sup>f</sup> | 5 | 0.2 | 0.2 | 0.1 |
| 15 | Keratoderma<sup>f</sup> | 42 | 1.4 | 1.7 | 0.9 | 42 | Verruca plana (flat warts)<sup>d</sup> | 5 | 0.2 | 0.2 | 0.1 |
| 16 | Tinea unguium (onychomycosis)<sup>a</sup> | 42 | 1.4 | 1.7 | 0.9 | 43 | Allergic dermatitis - n.f.d.<sup>e</sup> | 4 | 0.1 | 0.2 | 0.1 |
| 17 | Dermatitis – n.f.d.<sup>e</sup> | 38 | 1.3 | 1.5 | 0.8 | 44 | Molluscum contagiosum<sup>d</sup> | 4 | 0.1 | 0.2 | 0.1 |
| 18 | Intertrigo<sup>e</sup> | 38 | 1.3 | 1.5 | 0.8 | 45 | Prurigo nodularis<sup>e</sup> | 4 | 0.1 | 0.2 | 0.1 |
| 19 | Seborrhoeic dermatitis (seborrhoeic eczema)<sup>e</sup> | 36 | 1.2 | 1.4 | 0.8 | 46 | Verruca – n.f.d.<sup>d</sup> | 4 | 0.1 | 0.2 | 0.1 |
| 20 | Lichen simplex chronicus<sup>e</sup> | 27 | 0.9 | 1.1 | 0.6 | 47 | Idiopathische guttata hypomelanosus<sup>f</sup> | 3 | 0.1 | 0.1 | 0.1 |
| 21 | Photodermatitis<sup>e</sup> | 21 | 0.7 | 0.8 | 0.5 | 48 | Lipoma<sup>f</sup> | 3 | 0.1 | 0.1 | 0.1 |
| 22 | Fungal infection – n.f.d.<sup>a</sup> | 20 | 0.7 | 0.8 | 0.4 | 49 | Varices<sup>f</sup> | 3 | 0.1 | 0.1 | 0.1 |
| 23 | Tinea manuum<sup>a</sup> | 17 | 0.6 | 0.7 | 0.4 | 50 | Abscess<sup>c</sup> | 2 | 0.1 | 0.1 | 0.04 |
| 24 | Vitiligo<sup>f</sup> | 17 | 0.6 | 0.7 | 0.4 | 51 | Blistering disease – n.f.d.<sup>e</sup> | 2 | 0.1 | 0.1 | 0.04 |
| 25 | Wound(s)<sup>f</sup> | 17 | 0.6 | 0.7 | 0.4 | 52 | Chicken pox (varicella zoster)<sup>d</sup> | 2 | 0.1 | 0.1 | 0.04 |
| 26 | Vaginal candidiasis<sup>a</sup> | 16 | 0.5 | 0.6 | 0.4 | 53 | Dermatose – n.f.d.<sup>f</sup> | 2 | 0.1 | 0.1 | 0.04 |
| 27 | Onchocerciasis (river blindness)<sup>b*</sup> | 15 | 0.5 | 0.6 | 0.3 | 54 | Discoid lupus erythematosus<sup>e</sup> | 2 | 0.1 | 0.1 | 0.04 |
| 55 | Fixed drug eruption <sup>e</sup> | 2 | 0.1 | 0.1 | 0.04 | 79 | Lichen striatus <sup>e</sup> | 1 | 0.03 | 0.04 | 0.02 |
| 56 | Lymphatic filariasis <sup>b *</sup> | 2 | 0.1 | 0.1 | 0.04 | 80 | Lip licking dermatitis <sup>e</sup> | 1 | 0.03 | 0.04 | 0.02 |
| 57 | Naevus anaemicus <sup>f</sup> | 2 | 0.1 | 0.1 | 0.04 | 81 | Melasma <sup>f</sup> | 1 | 0.03 | 0.04 | 0.02 |
| 58 | Scar <sup>f</sup> | 2 | 0.1 | 0.1 | 0.04 | 82 | Naevus epidermalis <sup>f</sup> | 1 | 0.03 | 0.04 | 0.02 |
| 59 | Tinea incognito <sup>a</sup> | 2 | 0.1 | 0.1 | 0.04 | 83 | Nummular eczema (discoid eczema, - dermatitis) <sup>e</sup> | 1 | 0.03 | 0.04 | 0.02 |
| 60 | Verruca vulgaris (common wart) <sup>d</sup> | 2 | 0.1 | 0.1 | 0.04 | 84 | Papular eczema (papular dermatitis) <sup>e</sup> | 1 | 0.03 | 0.04 | 0.02 |
| 61 | Xerosis cuti (dry skin) <sup>f</sup> | 2 | 0.1 | 0.1 | 0.04 | 85 | Pemphigus vulgaris <sup>e</sup> | 1 | 0.03 | 0.04 | 0.02 |
| 62 | Acne keloidalis nuchae <sup>e</sup> | 1 | 0.03 | 0.04 | 0.02 | 86 | Pityriasis rosea <sup>e</sup> | 1 | 0.03 | 0.04 | 0.02 |
| 63 | Acrodermatitis <sup>e</sup> | 1 | 0.03 | 0.04 | 0.02 | 87 | Pyoderma gangrenosum ulcer <sup>c</sup> | 1 | 0.03 | 0.04 | 0.02 |
| 64 | Ano-genital herpes <sup>d</sup> | 1 | 0.03 | 0.04 | 0.02 | 88 | Scleroderma <sup>e</sup> | 1 | 0.03 | 0.04 | 0.02 |
| 65 | Aquagenic dermatitis <sup>e</sup> | 1 | 0.03 | 0.04 | 0.02 | 89 | Steroid related atrophy <sup>f</sup> | 1 | 0.03 | 0.04 | 0.02 |
| 66 | Auto-immune blistering disease – n.f.d. <sup>e</sup> | 1 | 0.03 | 0.04 | 0.02 | 90 | Steroid withdrawn dermatitis <sup>e</sup> | 1 | 0.03 | 0.04 | 0.02 |
| 67 | Chromoblastomycosis <sup>a *</sup> | 1 | 0.03 | 0.04 | 0.02 | 91 | Striae (stretch marks) <sup>f</sup> | 1 | 0.03 | 0.04 | 0.02 |
| 68 | Condylomata acuminata (ano-genital warts) <sup>d</sup> | 1 | 0.03 | 0.04 | 0.02 | 92 | Tuberous sclerosis <sup>f</sup> | 1 | 0.03 | 0.04 | 0.02 |
| 69 | Darier's disease <sup>e</sup> | 1 | 0.03 | 0.04 | 0.02 |  | **TOTAL** | **2,959** | **100.0** | **117.1** | **65.7** |
| 70 | Delusional parasitosis <sup>f</sup> | 1 | 0.03 | 0.04 | 0.02 |  | **\*NTDs (5 conditions)** | **280** | **9.5** | **11.1** | **6.2** |
| 71 | Diaper dermatitis (diaper rash) <sup>e</sup> | 1 | 0.03 | 0.04 | 0.02 | a | Primary fungal/yeast infections (11 conditions) | 1,675 | 56.6 | 66.3 | 37.2 |
| 72 | Eczema manuum (hand eczema, - dermatitis) <sup>e</sup> | 1 | 0.03 | 0.04 | 0.02 | b | Primary parasitic infections (3 conditions) | 234 | 7.9 | 9.3 | 5.2 |
| 73 | Erythema marginatum <sup>e</sup> | 1 | 0.03 | 0.04 | 0.02 | c | Primary bacterial infections (5 conditions) | 152 | 5.1 | 6.0 | 3.4 |
| 74 | Geographical tongue <sup>f</sup> | 1 | 0.03 | 0.04 | 0.02 | d | Primary viral infections (9 conditions) | 21 | 0.7 | 0.8 | 0.5 |
| 75 | Heat rash <sup>f</sup> | 1 | 0.03 | 0.04 | 0.02 | e | Other inflammatory diseases (36 conditions) | 697 | 23.6 | 27.6 | 15.5 |
| 76 | Herpes infection – n.f.d. <sup>d</sup> | 1 | 0.03 | 0.04 | 0.02 | f | Miscellaneous conditions (28 conditions) | 180 | 6.1 | 7.1 | 4.0 |
| 77 | Hypertrophic scar <sup>f</sup> | 1 | 0.03 | 0.04 | 0.02 | Abbreviations: n = number, n.f.d. = not further defined, NTDs = neglected tropical diseases. Co-infections other than impetigo (n=20) are not included in this list. |  |  |  |  |  |
| 78 | Kaposi sarcoma <sup>d</sup> | 1 | 0.03 | 0.04 | 0.02 |  |  |  |  |  |  |

Five different skin NTDs were seen: scabies, leprosy, onchocerciasis, lymphatic filariasis and chromoblastomycosis. These NTDs were found in 280 participants (9.5% of all diagnoses); none of the participants had more than 1 NTD. The top three skin NTDs seen were: scabies (n=217; 7.3% of all diagnoses), leprosy (n=45; 1.5% of all diagnoses) and onchocerciasis (n=15; 0.5% of all diagnoses) (Table 3). Scabies accounted for 77.5% of all identified NTDs, underscoring its dominant contribution to the overall NTD burden in these communities. Of the 45 persons affected by leprosy registered, 39 (0.9% of contacts screened) were newly diagnosed during the skin camps and started on multidrug therapy (MDT). One patient had previously been diagnosed with leprosy but had defaulted on treatment and was restarted on MDT. Five persons affected by leprosy had completed MDT but presented with visible leprosy-related disabilities. The already diagnosed patients were not among the index patients (n=44), who served as the entry points for the skin camps. If these index patients were also counted, a total of 89 (2.2%) persons affected by leprosy reside within the enrolled communities of 4,545 people.

Of the 44 patients of impetigo, 25 were recorded as second diagnosis which suggests co-infection. Secondary infection was also noted in 20 other patients, where ‘inflammation’ or ‘infection’ was recorded linked to forms of eczema, tinea capitis, tinea pedis, scabies, wounds, ulcers, and lichen simplex chronicus.

#### Skin disease per age group

Tinea capitis was significantly more common in children (*p*<0.001), with 666 (93.8%) of all 710 patients diagnosed in the 0–14-years group. In this group, tinea capitis accounted for 51.2% of all 1,302 skin disease diagnoses. Atopic dermatitis was also predominantly seen in children, with 57 diagnoses in the age group 0-14 years (*p*=0.014), representing 56.4% of all atopic dermatitis patients and 4.4% of all childhood diagnoses. Tinea corporis was also significantly more common in children, with 119 (58.3%) of all 204 patients in the 0–14-year group (*p*<0.001). Scabies was most frequently diagnosed among participants aged 0–14 and 15–29 years, accounting for 41.9% and 31.3% of all scabies patients, respectively (*p*<0.001). These correspond to 7.0% and 11.3% of all skin disease diagnoses within these age groups. Among adolescents and young adults (15–29 years), acne was the most frequent condition (*p*<0.001), comprising 96 (73.6%) of all 130 acne patients and 15.9% of all diagnoses within this age group. Pityriasis versicolor was also more prevalent in young adults (*p*< 0.001), with 226 (58.9%) of 384 patients diagnosed in the 15-44 year age range, representing 226 (20.9%) of all 1,080 diagnoses within these age groups. Tinea cruris showed a similar age distribution, predominating among adults aged 15–44 years 136 (63.8%) of 213 patients; *p*<0.001). In contrast, tinea pedis increased with age and was most often diagnosed in older adults (≥60 years), who accounted for 33.3% of all patients (*p*<0.001). When looking at the second most diagnosed NTD, leprosy, two of the newly detected leprosy patients were children below 15 years of age. The distribution of the ten most frequently diagnosed skin diseases across the age categories is summarised in Figure 2.

**Figure 2.**
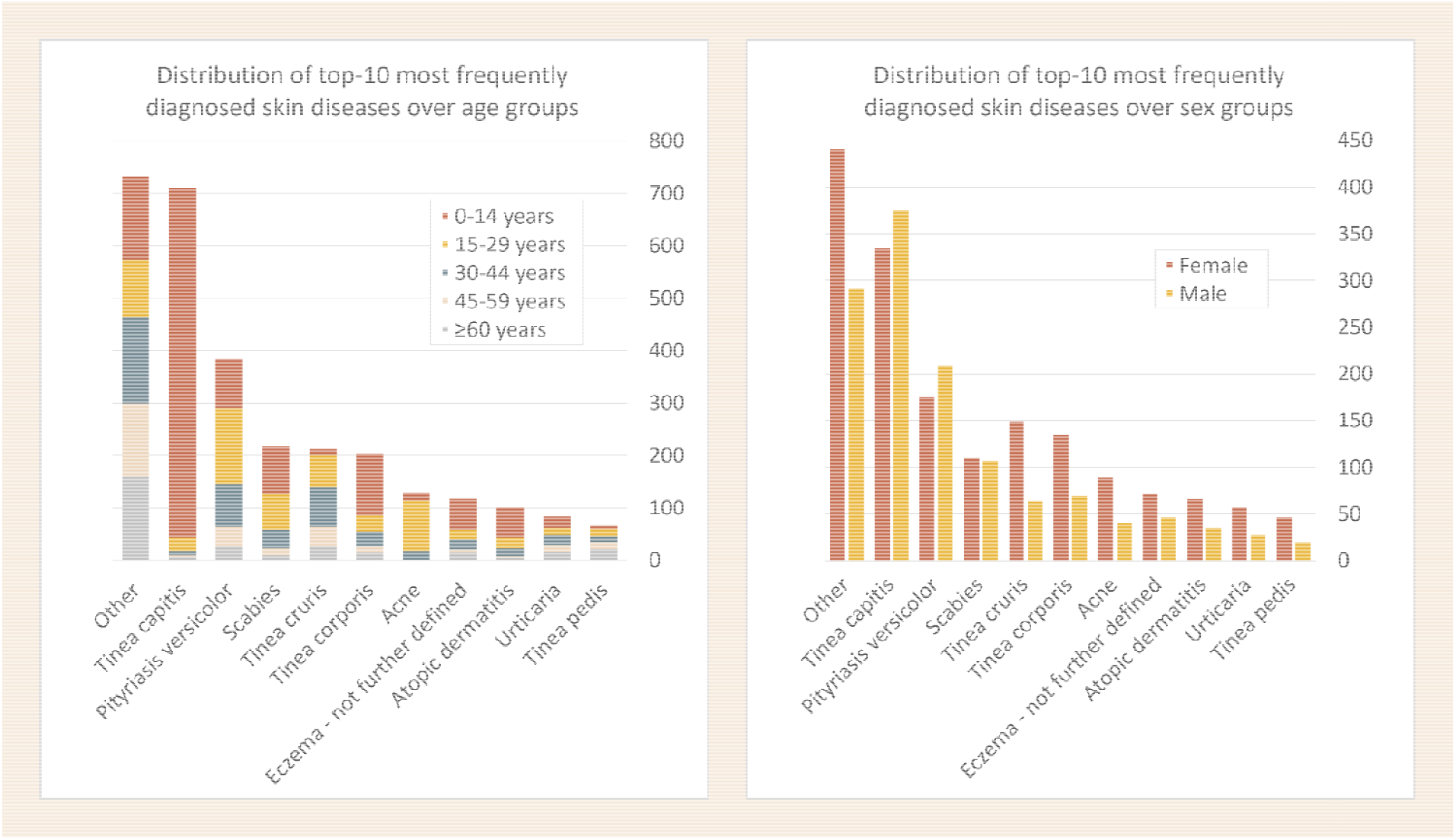
Distribution of the top-10 skin diseases detected across age and sex groups

### Skin diseases per sex

Of the 2,646 female participants screened, 1,435 (54.2%, 95%CI 52.3-56.1%) were diagnosed with one or more skin conditions while of the 1,855 male participants, 1,091 (58.8%, 95%CI 56.6-61.0%) had at least one skin condition, showing a slightly significant difference of 4.6%. Analysis of specific skin disease patterns revealed more prominent differences per sex (Figure 2). Male participants were more frequently diagnosed with tinea capitis, 375 (52.8%) of all 710 tinea capitis diagnoses were found in male participants, (*p*<0.001). Pityriasis versicolor was also found more often in male participants accounting for 209 (54.4%) of all 384 pityriasis versicolor diagnoses, (*p*<0.001). In contrast, female participants showed higher rates of tinea corporis accounting for 135 (66.2%) of all 204 tinea corporis diagnoses, (*p*=0.004). Women also accounted for 149 (70.0%) of all tinea cruris diagnoses, (*p*<0.001), 46 (69.7%) of all tinea pedis diagnoses, (*p*=0.030), and 57 (67.9%) of all urticaria diagnoses, (*p*=0.035).

### Treatment provided

In the PEP4LEP skin camps, only topical medication purchased from pharmaceutical suppliers and pharmacies in Tanzania was provided on site. When medication was unavailable, or when oral therapy was indicated, prescriptions were issued. Of the 2,527 participants diagnosed with skin diseases, 2,491 (98.6%) received one or more medications or emollients, either prescribed or provided directly at location. Antiparasitic agents, such as ivermectin and benzyl benzoate emulsion (BBE), were used for scabies. Albendazole was administered as an anthelmintic for suspected intestinal helminth infections in patients presenting with urticaria. Antibacterials, including fusidic acid and mupirocin creams, co-trimoxazole, and doxycycline, were prescribed for conditions such as folliculitis, impetigo, and secondary wound infections. Patients with leprosy received multidrug therapy (MDT) comprising rifampicin, clofazimine, and dapsone. Antifungal agents were among the most frequently dispensed medications, reflecting the high burden of fungal skin diseases. Clotrimazole and fluconazole creams, ketoconazole shampoo, terbinafine, and miconazole vaginal pessaries were used for dermatomycosis, (vaginal) candidiasis, and seborrhoeic dermatitis. Anti-inflammatory and corticosteroid treatments, hydrocortisone and betamethasone creams and prednisolone tablets, were prescribed for inflammatory dermatoses such as eczema, contact dermatitis, psoriasis, and lichen planus. The antiviral acyclovir was used for herpes infections, and cetirizine served as an antihistamine for conditions causing prurigo such as urticaria and allergic dermatitis. Keratolytics, for example benzoyl peroxide, salicylic acid ointment, and isotretinoin, were used for acne vulgaris and hyperkeratotic conditions including keratoderma. Antiseptics such as potassium permanganate external (PPE) solution and silver nitrate were applied for folliculitis and wound care, amongst others, while astringent or cauterizing agents including silver nitrate pencil and podophyllin solution were used to treat warts. Petroleum jelly was commonly applied as an emollient, and vitamin B complex was provided as a nutritional supplement for pellagra.

Referrals to health centres were made for severe or complex conditions, including blistering diseases, NTDs other than scabies, ulcers, or suspected chronic systemic infections such as HIV/AIDS. Participants also received targeted medical advice – for example, wearing shoes in case of leprosy or keratoderma, avoiding sunlight in photodermatitis, using compression stockings for varicose veins, discontinuing over-the-counter steroid creams in case of inappropriate usage, and receiving WASH-related guidance for infectious skin conditions. Reassurance and education for benign or self-limiting conditions was provided for diagnoses such as dermatosis papulosa nigra and idiopathic guttata hypomelanosis.

### Clinical presentation of sub-cohort

Among the contacts diagnosed with skin conditions at the skin camps, 1,841 persons (72.9%) were asked to participate in the additional survey on clinical presentation, with no refusals. For children who were too young to respond themselves, parents or caregivers provided the answers. Of this sub-sample, 817 (44.4%) were male and 1,024 (55.6%) were female. The majority (n=1,007, 54.7%) reported itchiness on their skin as their primary symptom at the moment of presentation. Among persons diagnosed with leprosy, the most frequently reported symptoms were numb skin lesions (n=10), followed by other skin changes (n=6), rash (n=4), ulcer (n=1), swelling (n= 1), and itching (n=1); none reported pain. The most affected body sites of participants detected with skin diseases who filled out the questionnaire were the scalp (n=503, 27.3%), followed by the trunk (n=293, 15.9%) and the face (n=268, 14.6%). A full list is available in Table 4. Among the 23 leprosy patients diagnosed in this sub-group, skin lesions were located on the trunk (n=9), the upper limbs (n=7), the face (n=6), and the lower limbs (n=1). The newly diagnosed leprosy patients did not exhibit lesions on the scalp, neck, genitals, groin, or gluteal region.

**Table 4.** Clinical presentation of the skin diseases (study sub-sample)

| Clinical presentation of 1,841 participants with skin conditions | Number of participants with skin diseases detected | % |
| --- | --- | --- |
| **Sex** |  |  |
| Female | 1,024 | 55.6 |
| Male | 817 | 44.4 |
| **Primary sign/symptom** |  |  |
| Itchiness | 1,007 | 54.7 |
| Skin rashes | 700 | 38.0 |
| Ulcer | 32 | 1.7 |
| Numbness on the lesions | 10 | 0.5 |
| Pain | 8 | 0.4 |
| Swelling | 2 | 0.1 |
| Fissures | 5 | 0.3 |
| Patches | 6 | 0.3 |
| Others (maceration, oozing, etc.) | 71 | 3.9 |
| **Primary body site affected** |  |  |
| Scalp | 503 | 27.3 |
| Trunk | 293 | 15.9 |
| Face | 268 | 14.6 |
| Genital, groin and gluteal region | 227 | 12.3 |
| Upper limbs | 201 | 10.9 |
| Lower limbs | 174 | 9.5 |
| Neck | 88 | 4.8 |
| Generalised | 60 | 3.3 |
| Others (interdigital space, underneath mammae, axilla) | 27 | 1.5 |
| **Primary skin lesion** |  |  |
| Papules | 860 | 46.7 |
| Patches/macules | 744 | 40.4 |
| Nodules | 22 | 1.2 |
| Blisters/vesicles/bullae | 20 | 1.1 |
| Ulcer | 22 | 1.2 |
| Swollen lower limb | 3 | 0.2 |
| Deformity | 6 | 0.3 |
| Plaque | 18 | 1.0 |
| Other (excoriations, maceration, etc.) | 146 | 7.9 |
| **Duration of the symptoms** |  |  |
| < 1 month | 395 | 21.5 |
| 1 month – 12 months | 935 | 50.8 |
| >12 months | 511 | 27.8 |
| **Family members with similar symptoms** |  |  |
| No | 1,425 | 77.4 |
| Yes | 415 | 22.5 |
| Missing value | 1 | 0.1 |
| **Seeking care prior to skin camp** |  |  |
| No | 1,321 | 71.8 |
| Yes | 520 | 28.3 |
| **Location of sought care prior to skin camp (n=520)** |  |  |
| Traditional healers | 10 | 1.9 |
| Health facility | 224 | 45.0 |
| Over-the-counter medication | 284 | 54.6 |
| Missing value | 2 | 0.4 |

The majority of the persons with skin diseases (n=935, 50.8%) reported having their lesions for more than one month but less than 12 months, while 511 (27.8%) reported experiencing their symptoms for more than a year. A total of 415 (22.5%) persons with skin diseases reported having family members who had similar complaints. A total of 520 (28.3%) of the 1,841 surveyed patients with a skin condition reported to have been looking for medical care prior to the skin camps (Table 4). Out of those 520 contacts, 284 (54.6%) reported having visited drug-selling shops or pharmacies, while 224 (43.2%) visited health facilities and 10 (1.9%) visited traditional healers.

## Discussion

This community-based study assessed the prevalence and characteristics of skin diseases among community contacts of leprosy index patients in three districts of Tanzania. More than half (56.1%) of the 4,501 participants were diagnosed with at least one dermatological condition, and over 11% of those affected had a skin NTD, underscoring a substantial burden of skin morbidity in these rural populations. Only a minority of participants with skin diseases surveyed had previously sought medical care, most of whom relied on pharmacies or informal drug sellers as their primary source of care.

### Skin diseases prevalence

The prevalence of skin diseases observed in this study exceeded earlier Tanzanian studies (27-35%) and was also higher than a more recent study screening schoolchildren from Côte d’Ivoire (34%), but was lower than reports from Cameroon (62%) and among Egyptian scholars (100%) [39,40]. Differences across studies likely reflect variations in diagnostic criteria, study populations, and access to dermatological care. For instance, the study from Egypt included benign naevi as diagnoses, inflating reported prevalence [41], while in this study naevi were only included when causing concern for study participants.

Multiple co-existing skin conditions were found in 15.6% of participants, which is lower than the school based study in Côte d’Ivoire where more than half of the children had multiple skin diseases, but higher than studies in Asia (3-9%) [40,42,43]. This co-occurrence may reflect a shared microbial aetiology, immune status, and age-specific disease patterns, as well as broader social determinants – such as hygiene, care access and overcrowding – that are closely linked to socioeconomic status [40,44].

The study population was predominantly young, with a median age of 20 years, slightly above the 2022 national median of 18 years [32]. The age-group distribution was broadly comparable to national patterns, with similar proportions in the 0-14 years (42.0% vs 42.8%), 30-44 years (15.9% vs 16.0%), and 45-59 years (10.7% vs 8.7%) groups, while our cohort included fewer persons aged 15-29 years (18.7% vs 26.9%) and a noticeably higher proportion aged 60 years and above (12.7% vs 5.7%). Participants diagnosed with skin diseases had a median age of 18 years, consistent with community-based studies in low- and middle-income countries [43,45,46]. Two of the three most common skin diseases – tinea capitis (94.7% of all affected participants were children; 49.7% of all paediatric skin disease diagnoses) and scabies (46.5% of affected participants were children; 7.5% of paediatric diagnoses) – showed a clear predominance in childhood, which reflects the vulnerability of younger age groups in resource-limited environments [47–49]. Reviews show that scabies affects 5-50% of children in resource-poor tropical regions, and that fungal skin diseases are highly prevalent among children in sub-Saharan Africa, with the pooled paediatric prevalence of tinea capitis estimated at 23% across several African countries.

Males showed a slightly higher prevalence of skin diseases (58.8%) than females (54.2%), which may reflect differences in occupational and climatic exposure, hygiene practices, or biological factors [50,51]. A school-based study in Côte d’Ivoire also reported a slightly higher skin disease prevalence among males, though the sex distribution varied by disease [40]. In both studies, tinea capitis was more frequent in males; however, in Côte d’Ivoire, tinea pedis was also more common in males and pityriasis versicolor predominated in females, contrasting with the Tanzanian findings. Studies from India, Egypt and Bangladesh likewise reported lower prevalence rates among females, but lower (20–38%) than those observed in this study [43,52–54].

When combined, fungal and yeast infections affected over half of participants, with tinea capitis being the most frequent diagnosis accounting for 15.8%. While the global prevalence of superficial fungal infections is estimated at around 10% [55], rates are considerably higher in low- and middle-income settings [28,29,40,53,54,56]. A literature review by Rodríguez-Cerdeira et al. reported that tinea capitis frequencies in African countries range from 0.4% to 88% [57]. Risk factors for fungal infections include warm and humid climates, as well as close animal contact, overcrowding, inadequate WASH facilities, and limited healthcare access – the latter often linked to poverty. Comorbidities such as HIV/AIDS, kidney failure, and diabetes may further increase susceptibility. The 2022–2023 Tanzania HIV Impact Survey reported adult HIV prevalences of 3.3% in Morogoro and 2.6% in Lindi, both below the mainland average of 4.5% [58]. Although national incidence has declined [59], reductions in global health funding programmes, such as seen in the *President’s Emergency Plan for AIDS Relief* (PEPFAR), may reverse progress and indirectly increase dermatological complications associated with HIV [60].

### Bacterial co-infections and WASH

Bacterial co-infections linked to compromised skin integrity, for example inflamed eczema, tinea capitis, scabies, and ulcers, were not routinely recorded as separate secondary diagnoses unless clearly identified (e.g. impetigo). However, all affected patients received appropriate treatment, when available, or prescriptions. Although most bacterial skin infections resolve without major complications, some can lead to severe, sometimes even fatal, outcomes such as sepsis and nephritis, particularly among immunocompromised persons [61,62]. The high occurrence of fungal, bacterial, and parasitic infections in this study highlights the influence of socioeconomic factors, particularly limited access to hygiene [63–65]. In 2022, the World Bank highlighted that inadequate WASH services continue to pose major health and economic challenges in Tanzania [66]. Although gradual progress has been made, only about 64% of households had access to basic water supply and 48% to basic hygiene facilities. Coverage remains particularly low in rural areas, such as our study settings. Limited WASH access sustains the transmission of common skin infections and other diseases. The health impact of inadequate WASH contributes to the loss of millions of working days due to related illnesses and exacerbates social inequalities, particularly among women [15,66,67]. Skin camps also serve as an opportunity to provide targeted health education on hygiene and WASH practices, which may help reduce the risk of recurrent or preventable skin infections. In line with WHO’s NTD Road Map, improved coordination between WASH and (skin) NTD programmes could maximise health impact and cost-effectiveness. Improving WASH infrastructure, alongside health education and expanded access to dermatological services, is crucial to reduce the burden of preventable skin diseases in resource-limited settings [20].

### Skin NTDs

Skin NTDs were identified in 6.2% of all participants (11.1% of participants with skin diseases) accounting for 9.5% of dermatological diagnoses, with scabies being the most common, followed by leprosy. This percentage of 6.2% exceeds findings from Côte d’Ivoire, where just over 1% of schoolchildren aged 5 to 15 years screened had a skin NTD (scabies or leprosy) [40]; however, the present study included community members of all ages, which may explain the difference. When only looking at children in the same age group in this Tanzanian study, scabies and leprosy were also the only NTDs detected. Two of the leprosy patients newly detected in Tanzania were below 15 years of age, which is a sign of recent *Mycobacterium leprae* infection and thus ongoing transmission in their communities [68]. Other population-based data from comparable settings remain scarce, but these Tanzanian findings align with WHO estimates that skin NTDs account for up to 10% of the dermatological burden in tropical and resource-limited settings [4]. Five skin NTDs were detected in this Tanzanian cohort – scabies, leprosy, onchocerciasis, lymphatic filariasis, and chromoblastomycosis – demonstrating the feasibility of integrated, community-based skin screening that targets both common skin diseases and skin NTDs within one intervention. These results line up with WHO data showing that 37 of the 47 countries in the WHO African Region are co-endemic for five or more NTDs [69], and support WHO’s recommendation to adopt integrated strategies for diagnosis and care [19,20,70]. Scabies, affecting an estimated 200 million people globally, is strongly associated with poverty, overcrowding, and limited hygiene and healthcare access [48]. Approximately one-third of scabies patients develop bacterial superinfection [71], reinforcing the importance of WASH facilities, hygiene education and available household-level treatment. Study findings were shared with Tanzania’s Ministry of Health to support the planning of targeted NTD interventions. For example, the relatively high number of scabies diagnoses found in several communities may warrant mass drug administration (MDA) with ivermectin, in line with WHO guidance for areas where prevalence exceeds 10% [48]. Ivermectin-based MDA for onchocerciasis is already ongoing in endemic settings in the study districts [20]. In regions co-endemic for onchocerciasis and lymphatic filariasis, albendazole is advised to be co-administered with ivermectin during community-based treatment. Combining integrated skin screening and single-dose rifampicin (SDR-PEP) for leprosy prevention with MDA campaigns for other NTDs in endemic settings – such as in communities covered by this study – could enhance programme efficiency and cost-effectiveness. This co-endemicity-driven, integrated approach aligns with WHO’s vision for cross-cutting, resource-optimised NTD control strategies. Future research on combined preventive interventions is advised.

### Addressing dermatological workforce shortages

The national shortage of dermatologists in Tanzania – only 48 dermatovenereologists for a population exceeding 60 million – highlights the urgent need for increased specialist training opportunities, as well as task-sharing and capacity strengthening among primary and community healthcare workers [3,15,19,20,66,71,72]. Given that more than 3,000 dermatological conditions are recognised, and only a limited number can realistically be taught within the constrained training time available to non-dermatologists, prioritised and context-specific training becomes essential [73,74]. Strengthening the capacity of primary healthcare workers to recognise and manage common, high-prevalence skin conditions – alongside less frequent but clinically severe or time-sensitive presentations such as skin NTDs, blistering disorders, extensive bacterial infections, (potential) oncological conditions, and HIV-related skin diseases – would address much of the skin-disease burden. Targeted training focused on the 10-30 most common skin diseases – leaning towards the lower end when conditions are grouped (e.g., fungal infections) – alongside the timely recognition of clinically severe, time-sensitive, or high-risk presentations beyond this ‘top 30’ would provide a strong foundation for frontline dermatological care. This is especially relevant as all of these higher-morbidity conditions named above were observed in this study.

Other skin conditions can then be managed through referral pathways for further expertise and diagnostics, or supported via tele-dermatology and mobile health (mHealth) tools, including applications using artificial intelligence (AI) [14,15,25,75]. A clearer definition of the most frequent and high-priority conditions as provided in this study can also inform machine-learning training datasets, helping ensure that AI tools are better adapted to the epidemiology and diagnostic needs of low-income settings.

In addition, skin camps can function as practical training sites where primary healthcare workers gain hands-on exposure to a broad spectrum of skin conditions and receive real-time coaching from dermatologically trained staff, enabled by the high number and diversity of skin conditions seen [30,76,77].

### Healthcare-seeking behaviour

Less than one-third of participants with skin conditions had previously sought care, most relying on pharmacies or informal drug sellers. This reliance on often unregulated providers may delay accurate diagnosis, foster complications – such as the steroid-induced dermatoses observed in two patients in this cohort – and contribute to antimicrobial misuse and resistance. Limited awareness and stigma surrounding dermatological diseases further discourage patients from seeking care [3,14,15,44,78,79].

### Strengths and limitations

This study is among the few recent studies to provide community-based dermatological data sub-Sahara Africa, offering a more representative perspective than school- or clinic-based surveys [7,26–29,39,40,42,80,81]. The absence of routine surveillance leads to underestimation of the true burden of skin diseases, particularly in hard-to-reach communities where skin NTDs remain largely underreported [15,19]. Integrated community-based dermatological screening generates essential evidence for planning and monitoring skin health interventions and supports the WHO’s global strategy for integrated skin NTD control [19,20,70]. More results from the PEP4LEP and PEP4LEP 2.0 study, also including data from Ethiopia and Mozambique are expected soon [30].

Including only dermatologist-led skin camps ensured diagnostic reliability. Given the profound shortage of dermatologists in Tanzania, the study provided a valuable and unusual opportunity to capture community-level data [24,72]. Not surprisingly, other studies have confirmed that dermatologists achieve higher diagnostic accuracy of skin conditions than general or specialized clinicians [82,83]. Histopathological examination was not undertaken due to feasibility constraints in the community setting, and this also reflecting routine practice in Tanzania and many other low-resource contexts where access to such testing is often limited [3,14,75]. Participants requiring additional evaluation were referred to health centres through established pathways, consistent with implementation research approaches [75,84]. Tran et al. similarly used dermatologists’ diagnoses as a reference standard in the absence of histology in an Australian study [82].

Some participants presented with anogenital conditions, including vaginal candidiasis, herpes (vesicles), condylomata acuminata, and pruritus scroti. These were addressed within the scope of dermato-venereology when symptoms were reported, although the cohort was not comprehensively screened for anogenital or sexually transmitted infections.

Strictly categorising skin diseases can be challenging, and variations between studies are common [3,6,14,40,46,80], as many conditions have mixed or secondary aetiologies. For example, angular cheilitis, intertrigo, and diaper dermatitis frequently show fungal colonisation but are primarily driven by irritant dermatitis. In addition, acne largely stems from excess sebum causing blocked sebaceous ducts, which may progress to inflammatory lesions [9]. In this study, classification was based on the predominant aetiology, broadly following the system proposed by Yotsu et al. [40].

The World Health Organization Resolution WHA78.15 states that “… the majority of the skin-disease burden in any community is caused by about 10 common general skin diagnoses …” and emphasises that most patients with these diagnoses can be managed at primary care level with essential medicines, the right training and support [15]. However, the resolution does not specify which ten conditions these are. In this cohort, the ten most frequent skin disease diagnoses (10 out of 92; 10.9%) accounted for 75.3% of all recorded diagnoses and 88.1% of all participants diagnosed with skin conditions. The Global Burden of Disease Study 2021 identified 12 major skin diseases and categories – dermatitis, psoriasis, bacterial skin diseases, scabies, fungal skin diseases, viral skin diseases, acne vulgaris, alopecia areata, pruritus, urticaria, bedsores (decubitus ulcer), and other skin and subcutaneous diseases – which closely overlap with the conditions observed in this study [6]. Improving surveillance and standardising data collection would help clarify the most prevalent conditions and refine priority areas for contextual primary care training [14,15].

Expanding community-based dermatological services, strengthening health worker training, and providing targeted health education through the PEP4LEP project and the follow-up PEP4LEP 2.0 are likely to help reduce inequities in health service coverage [15,19,20,30,67,77]. Despite the high turnout at the skin camps overall, individuals with visible or symptomatic skin conditions may have been more likely to attend, although the availability of leprosy preventive medication may also have encouraged participation among non-symptomatic community members. This aspect will be explored in greater depth through the qualitative data collected in PEP4LEP and PEP4LEP 2.0 [30].

While this study was conducted in Tanzania, its findings are relevant to comparable resource-limited settings across sub-Saharan Africa and beyond. The diagnostic spectrum observed and the demonstrated feasibility of integrated skin screening suggest that similar community-based approaches could be replicated in other regions where dermatological care remains limited. This also includes the combined approach of integrated skin screening with SDR-PEP for leprosy prevention, which proved feasible within the community setting [30,31]. Besides their role in screening, improving care access, and serving as practical training sites [30,76,77], skin camps can also help identify service needs – including medication availability – within local health systems, providing useful insights for policymakers. Integrated community skin screening, as illustrated in this study, can contribute to the broader goals of universal health coverage and aligns with the WHA’s global call to prioritise skin health [15]. Sustained progress in global dermatological care, including for skin NTDs, will depend on (technical) innovation, strong political and financial commitment, intersectoral collaboration, and active community engagement to ensure equitable, long-term impact and make skin health accessible for all [13– 15,65,67,69].

## Conclusion

This study provides one of the few recent community-based dermatological datasets from sub-Saharan Africa, offering essential evidence on the burden of skin diseases and skin-related NTDs in resource-limited settings. More than half of all screened participants were diagnosed with one or more skin conditions. Five NTDs were identified, affecting 6.2% of all participants and accounting for nearly one in ten skin disease diagnoses made. These findings highlight the public health importance of community-based integrated dermatological care through skin camps and the feasibility of combining screening for common skin diseases and NTDs with leprosy chemoprophylaxis (SDR-PEP) in endemic regions where specialised care is scarce. This aligns with the recent World Health Assembly resolution recognising skin diseases as a global health priority and promoting community-based integrated dermatological care [15]. Expanding the quality of service delivery within such models will depend on enhanced health worker training and supervision, strong community engagement, effective surveillance, and reliable logistics that ensure the availability of necessary care. Sustainable financing and strong political and stakeholder commitment are essential to expand dermatological care in community settings that currently lack access, and, in turn, to reduce preventable morbidity and microbial transmission in Tanzania and comparable settings worldwide.

## Data Availability

The original data set generated and analysed during the current study will be stored for a period of 25 years. The anonymised, relevant raw dataset is available for authorised reuse, such as future analyses or replication, via the international NTD information platform infoNTD.org.

https://www.leprosy-information.org/resource/pep4lep-20-raw-data-depository

## Acknowledgments

The authors extend their sincere gratitude to all study participants who voluntarily took part, and to the health workers who supported the screening and the running of the skin camps. We are grateful to the full research consortium, including all researchers and committee members – with special thanks to Marente Mol, Isabela De Caux Bueno, Sophie Vissers, Benita Jansen, Thomas Hambridge, David Blok, Colette van Hees, Carolin Gunesch, Kidist Bobosha, Paul Saunderson – along with all other project staff and interns for their valuable contributions. We also acknowledge our funders, the European and Developing Countries Clinical Trials Partnership (EDCTP), its contributors and the Leprosy Research Initiative (LRI), whose support made this work possible.

## Funding

This work is part of the PEP4LEP and PEP4LEP 2.0 projects. PEP4LEP is part of the EDCTP2 programme supported by the European Union awarded to NLR/LM (grant number RIA2017NIM-1839-PEP4LEP), and the Leprosy Research Initiative (LRI; www.leprosyresearch.org) awarded to NLR/LM (grant number 707.19.58.).The PEP4LEP 2.0 project is part of the Global Health EDCTP3 Joint Undertaking, co-funded by the European Union (grant number 101145677-PEP4LEP2.0). Views and opinions expressed are however those of the authors only. The funders had no role in study design, data collection, and analysis, the decision to publish, or the preparation of the manuscript.

## Contributions

All co-authors are involved in the PEP4LEP and PEP4LEP 2.0 studies. The PEP4LEP proposal, protocol and/or data collection forms were created by AS, RvW, LM, JHR, CK. NM diagnosed the skin diseases during the skin camps as dermatologist, and designed and conducted the additional survey. PN entered the data into the REDCap database. NM, RvW and AS analysed the data. AS and NM drafted the manuscript together with RvW and LM. All co-authors reviewed the draft manuscript and provided input as needed. AI technology (ChatGPT.com, developed by OpenAI) was used for spelling checks and to improve readability. The manuscript was also reviewed by the PEP4LEP 2.0 International Publication Committee according to study guidelines [30].

## Data sharing statement

The original data set generated and analysed during the current study will be stored for a period of 25 years [30]. The anonymised, relevant raw dataset is available for authorised reuse, such as future analyses or replication, via the international NTD information platform: www.infoNTD.org [85].

## Conflict of interest

Authors declare no conflict of interest.

## Notes

### Competing Interest Statement

The authors have declared no competing interest.

### Clinical Trial

PACTR202303742093429

### Clinical Protocols

https://pubmed.ncbi.nlm.nih.gov/34446483/

https://nlrinternational.org/what-we-do/projects/https-nlrinternational-trybes-work-what-we-do-projects-pep4lep/

### Author Declarations

Permission to conduct the study was approved by the joint CUHAS/BMC Research Ethics and Review Committee (clearance no. CREC/364/2019). The PEP4LEP project in Tanzania was approved by the National Institute for Medical Research (NIMR) of Tanzania (reference number; NIMR/HQ/R.8c/Vol.1/1530).

## References

1 Hay RJ, Johns NE, Williams HC, et al. The Global Burden of Skin Disease in 2010: An Analysis of the Prevalence and Impact of Skin Conditions. J Invest Dermatol. 2014;134:1527–34. doi: 10.1038/jid.2013.446

2 Ryan TJ, Ersser SJ, Fuller LC. The Public Health Intervention of Skin Care for All: Community Dermatology. In: Maddock J, ed. Public Health - Social and Behavioral Health. InTech 2012. https://www.intechopen.com/chapters/36962 (accessed 25 November 2025)

3 Seth D, Cheldize K, Brown D, et al. Global Burden of Skin Disease: Inequities and Innovations. Curr Dermatol Rep. 2017;6:204–10. doi: 10.1007/s13671-017-0192-7

4 World Health Organization. WHO’s first global meeting on skin NTDs calls for greater efforts to address their burden. 2023. https://www.who.int/news/item/31-03-2023-who-first-global-meeting-on-skin-ntds-calls-for-greater-efforts-to-address-their-burden (accessed 24 August 2025)

5 Giesey RL, Mehrmal S, Uppal P, et al. Dermatoses of the world: Burden of skin disease and associated socioeconomic status in the world. J Am Acad Dermatol. 2021;84:556–9. doi: 10.1016/j.jaad.2020.05.157

6 Huai P, Xing P, Yang Y, et al. Global burden of skin and subcutaneous diseases: an update from the Global Burden of Disease Study 2021. Br J Dermatol. 2025;192:1136–8. doi: 10.1093/BJD/LJAF071

7 Onayemi O, Isezuo SA, Njoku CH. Prevalence of different skin conditions in an outpatients’ setting in north-western Nigeria. Int J Dermatol. 2005;44:7–11. doi: 10.1111/J.1365-4632.2004.02298.x

8 Gibbs S. Skin disease and socioeconomic conditions in rural Africa - Tanzania. Int J Dermatol. 1996;35:633–9. doi: 10.1111/j.1365-4362.1996.tb03687.x

9 van Hees Colette, Naafs Ben. Common Skin Diseases In Africa: An Illustrated Guide. 2004. https://www.medbox.org/document/common-skin-diseases-in-africa-an-illustrated-guide (accessed 25 August 2025)

10 Chandler DJ, Fuller LC. The Skin-A Common Pathway for Integrating Diagnosis and Management of NTDs. Trop Med Infect Dis. 2018;3:101. doi: 10.3390/tropicalmed3030101

11 Yotsu RR, Fuller LC, Murdoch ME, et al. A global call for action to tackle skin-related neglected tropical diseases (skin NTDs) through integration: An ambitious step change. PLoS Negl Trop Dis. 2023;17:e0011357. doi: 10.1371/JOURNAL.PNTD.0011357

12 World Health Organization. Skin-related neglected tropical diseases. https://www.who.int/teams/control-of-neglected-tropical-diseases/interventions/strategies/skin-ntds (accessed 22 August 2025)

13 Yotsu RR, Coffie Comoé C, Taïba Ainyakou G, et al. Impact of common skin diseases on children in rural Côte d’Ivoire with leprosy and Buruli ulcer co-endemicity: A mixed methods study. PLoS Negl Trop Dis. 2020;14:e0008291. doi: 10.1371/journal.pntd.0008291

14 Mugisha N, Ghanem L, Komi OAI, et al. The screening and management of skin diseases in remote African regions: a narrative review. Postgrad Med J. 2025;101:504–10. doi: 10.1093/POSTMJ/QGAE133

15 World Health Organization. World Health Assembly (WHA) Resolution - Skin diseases as a global public health priority. 2025. https://apps.who.int/gb/ebwha/pdf_files/EB156/B156_(24)-en.pdf (accessed 30 October 2025)

16 World Health Organization. Recognizing neglected tropical diseases through changes on the skin. 2018. https://www.who.int/publications/i/item/9789241513531 (accessed 25 November 2025)

17 Hay RJ, Asiedu K. Skin-Related Neglected Tropical Diseases (Skin NTDs)—A New Challenge. Trop Med Infect Dis. 2018;4:4. doi: 10.3390/tropicalmed4010004

18 Yotsu RR. Integrated Management of Skin NTDs-Lessons Learned from Existing Practice and Field Research. Trop Med Infect Dis. 2018;3:120. doi: 10.3390/tropicalmed3040120

19 World Health Organization. Ending the neglect to attain the Sustainable Development Goals: A strategic framework for integrated control and management of skin-related neglected tropical diseases. 2022. https://www.who.int/publications/i/item/9789240051423 (accessed 24 August 2025)

20 World Health Organization. Ending the neglect to attain the Sustainable Development Goals: A road map for neglected tropical diseases 2021–2030. 2021. https://www.who.int/publications/i/item/9789240010352 (accessed 25 November 2025)

21 Alchorne MM de A, Conceição K da C, Barraza LL, et al. Dermatology in black skin. An Bras Dermatol. 2024;99:327–41. doi: 10.1016/J.ABD.2023.10.001

22 Brown-Korsah JB, McKenzie S, Omar D, et al. Variations in genetics, biology, and phenotype of cutaneous disorders in skin of color – Part I: Genetic, biologic, and structural differences in skin of color. J Am Acad Dermatol. 2022;87:1239–58. doi: 10.1016/j.jaad.2022.06.1193

23 Malone Mukwende, Tamony Peter, Turner Margot. Mind the Gap: A handbook of clinical signs in Black and Brown skin. 1st ed. London: St. Georges University of London 2020. https://www.blackandbrownskin.co.uk/mindthegap (accessed 25 August 2025)

24 Mosam A, Todd G. Dermatology Training in Africa: Successes and Challenges. Dermatol Clin. 2021;39:57–71. doi: 10.1016/j.det.2020.08.006

25 Groh M, Badri O, Daneshjou R, et al. Deep learning-aided decision support for diagnosis of skin disease across skin tones. Nat Med. 2024;30:573–83. doi: 10.1038/s41591-023-02728-3

26 Ayanlowo O, Puddicombe O, Gold-Olufadi S. Pattern of skin diseases amongst children attending a dermatology clinic in Lagos, Nigeria. Pan Afr Med J. 2018;29:1–10. doi: 10.4314/pamj.v29i1.

27 Kiprono SK, Muchunu JW, Masenga JE. Skin diseases in pediatric patients attending a tertiary dermatology hospital in Northern Tanzania: a cross-sectional study. BMC Dermatol. 2015;15:16. doi: 10.1186/s12895-015-0035-9

28 Khan SS, Ashcroft DM, Dlova N, et al. A skin disease survey of the Maasai in North Eastern Tanzania. Br J Dermatol. 2023;188:303–4. doi: 10.1093/bjd/ljac065

29 Satimia FT, McBride SR, Leppard B. Prevalence of skin disease in rural Tanzania and factors influencing the choice of health care, modern or traditional. Arch Dermatol. 1998;134:1363–6. doi: 10.1001/ARCHDERM.134.11.1363,

30 Schoenmakers A, Hambridge T, van Wijk R, et al. PEP4LEP study protocol: integrated skin screening and SDR-PEP administration for leprosy prevention: comparing the effectiveness and feasibility of a community-based intervention to a health centre-based intervention in Ethiopia, Mozambique and Tanzania. BMJ Open. 2021;11:e046125. doi: 10.1136/BMJOPEN-2020-046125

31 Hambridge T, Blok DJ, Mamo E, et al. The Epidemiological Impact of Community-Based Skin Camps on Leprosy Control in East Hararghe Zone, Ethiopia: a Modelling Study. J Epidemiol Glob Health. 2025;15:1–7. doi: 10.1007/S44197-025-00370-5

32 Ministry of Finance and Planning Tanzania, National Bureau of Statistics, President’s Office, et al. The 2022 Population and Housing Census: Age and Sex Distribution Report Tanzania Mainland. 2022. https://sensa.nbs.go.tz/publication/volume2b.pdf (accessed 16 September 2025)

33 The United Republic of Tanzania (URT), Ministry of Finance, Tanzania National Bureau of Statistics, et al. The 2022 Population and Housing Census: Morogoro Region Basic Demographic and Socio-Economic Profile Report; Tanzania. 2024. https://sensa.nbs.go.tz/publication/Morogoro.pdf (accessed 16 September 2025)

34 Ellen F Ter, Tielens K, Fenenga C, et al. Implementation approaches for leprosy prevention with single-dose rifampicin: A support tool for decision making. PLoS Negl Trop Dis. 2022;16. doi: 10.1371/JOURNAL.PNTD.0010792

35 World Health Organization. Guidelines for the diagnosis, treatment and prevention of leprosy. 2018. https://www.who.int/publications/i/item/9789290226383 (accessed 25 November 2025)

36 World Health Organization. Leprosy/Hansen disease: Contact tracing and post-exposure prophylaxis. 2020. https://www.who.int/publications/i/item/9789290228073 (accessed 25 November 2025)

37 Vanderbilt University. About – REDCap. https://projectredcap.org/about/ (accessed 25 November 2025)

38 Pan African Clinical Trials Registry (PACTR). Pan African Clinical Trials Registry. 2023. https://pactr.samrc.ac.za/TrialDisplay.aspx?TrialID=24385 (accessed 25 November 2025)

39 Mahé A, Prual A, Konaté M, et al. Skin diseases of children in Mali: a public health problem. Trans R Soc Trop Med Hyg. 1995;89:467–70. doi: 10.1016/0035-9203(95)90068-3

40 Yotsu RR, Kouadio K, Vagamon B, et al. Skin disease prevalence study in schoolchildren in rural Côte d’Ivoire: Implications for integration of neglected skin diseases (skin NTDs). PLoS Negl Trop Dis. 2018;12:e0006489. doi: 10.1371/journal.pntd.0006489

41 El-Khateeb EA, Lotfi RA, Abdel-Aziz KM, et al. Prevalences of skin diseases among primary schoolchildren in Damietta, Egypt. Int J Dermatol. 2014;53:609–16. doi: 10.1111/ijd.12335

42 Dogra S, Kumar B. Epidemiology of Skin Diseases in School Children: A Study from Northern India. Pediatr Dermatol. 2003;20:470–3. doi: 10.1111/j.1525-1470.2003.20602.x

43 Ahmed N, Islam M, Farjana S. Pattern of Skin Diseases: Experience from a Rural Community of Bangladesh. Bangladesh Med J. 2014;41:50–2. doi: 10.3329/bmj.v41i1.18784

44 Inanir I, Şahin MT, Gündüz K, et al. Prevalence of Skin Conditions in Primary School Children in Turkey: Differences Based on Socioeconomic Factors. Pediatr Dermatol. 2002;19:307–11. doi: 10.1046/j.1525-1470.2002.00087.x

45 Bommakanti J, Pendyala P. Pattern of skin diseases in rural population: a cross sectional study at Medchal mandal, Rangareddy district, Telangana, India. Int J Res Med Sci. Published Online First: 2017. doi: 10.18203/2320-6012.ijrms20164443

46 Shrestha R, Shrestha DP, Lama L, et al. Pattern of skin diseases in a rural village development community of Nepal. Nepal J Dermatol Venereol Leprol. 2014;12:41–4. doi: 10.3126/njdvl.v12i1.10595

47 Liu X, Zhang Y, Hong Y, et al. Global burden of fungal skin diseases: An update from the Global Burden of Diseases Study 2019. Mycoses. 2024;67. doi: 10.1111/MYC.13770,

48 World Health Organization. Scabies. 2023. https://www.who.int/news-room/fact-sheets/detail/scabies (accessed 22 August 2025)

49 Bongomin F, Olum R, Nsenga L, et al. Estimation of the burden of tinea capitis among children in Africa. Mycoses. 2021;64:349–63. doi: 10.1111/MYC.13221

50 Dao H, Kazin RA. Gender differences in skin: A review of the literature. Gend Med. 2007;4:308–28. doi: 10.1016/S1550-8579(07)80061-1

51 Lagacé F, D’Aguanno K, Prosty C, et al. The Role of Sex and Gender in Dermatology - From Pathogenesis to Clinical Implications. J Cutan Med Surg. 2023;27:NP1–36. doi: 10.1177/12034754231177582

52 Abdel-Hafez K, Abdel-Aty MA, Hofny ERM. Prevalence of skin diseases in rural areas of Assiut Governorate, Upper Egypt. Int J Dermatol. 2003;42:887–92. doi: 10.1046/j.1365-4362.2003.01936.x

53 Grover S, Ranyal RK, Bedi MK. A cross section of skin diseases in rural Allahabad. Indian J Dermatol. 2008;53:179– 81. doi: 10.4103/0019-5154.44789

54 Singhal R, Talati K, Gandhi B, et al. Prevalence and pattern of skin diseases in tribal villages of Gujarat: A teledermatology approach. Indian J Community Med. 2020;45:199. doi: 10.4103/ijcm.IJCM_76_19

55 Urban K, Chu S, Giesey RL, et al. Burden of skin disease and associated socioeconomic status in Asia: A cross-sectional analysis from the Global Burden of Disease Study 1990-2017. JAAD Int. 2021;2:40–50. doi: 10.1016/j.jdin.2020.10.006

56 Walker SL, Shah M, Hubbard VG, et al. Skin disease is common in rural Nepal: results of a point prevalence study. British Journal of Dermatology. 2007;0:070818011041017 doi: 10.1111/j.1365-2133.2007.08107.x

57 Rodríguez-Cerdeira C, Martínez-Herrera E, Szepietowski JC, et al. A systematic review of worldwide data on tinea capitis: analysis of the last 20 years. J Eur Acad Dermatol Venereol. 2021;35:844–83. doi: 10.1111/jdv.16951

58 National Bureau of Statistics United Republic of Tanzania. Summary Sheet | December 2023 - The Tanzania HIV Impact Survey 2022-2023. 2023. https://nbs.go.tz/nbs/takwimu/THIS2022-2023/THIS2022-2023_Summary_Sheet.pdf?utm (accessed 24 October 2025)

59 Unaids. UNAIDS data 2024. 2024. http://www.wipo.int/amc/en/mediation/rules (accessed 24 October 2025)

60 Hontelez JAC, Goymann H, Berhane Y, et al. The impact of the PEPFAR funding freeze on HIV deaths and infections: a mathematical modelling study of seven countries in sub-Saharan Africa. EClinicalMedicine. 2025;83:103233. doi: 10.1016/j.eclinm.2025.103233

61 Flohr C, Hay R. Putting the burden of skin diseases on the global map. Br J Dermatol. 2021;184:189–90. doi: 10.1111/BJD.19704

62 Duncan N, Whittaker L, Oakley A, et al. Bacterial skin infections. 2023. https://dermnetnz.org/topics/bacterial-skin-infections (accessed 4 November 2025)

63 Aman B, Taj M, Samreen Z, et al. Skin infection risk factors and its management. J Biodivers Environ Sci. 2020;16:117–33.

64 Pathak R, Shrestha S, Poudel P, et al. Association of Socio-Demographic Factors and Personal Hygiene with Infectious Childhood Dermatoses. Skin Health Dis. 2023;3. doi: 10.1002/SKI2.219

65 Mugisha N, Ghanem L, Komi OAI, et al. The screening and management of skin diseases in remote African regions: a narrative review. Postgrad Med J. 2025;101:504–10. doi: 10.1093/POSTMJ/QGAE133

66 World Bank. Tanzania Economic Update - Clean Water, Bright Future[]: The Transformative Impact of Investing in WASH (English). Tanzania economic update; issue no. 18. Washington, D.C. 2022. https://documents.worldbank.org/en/publication/documents-reports/documentdetail/099141002082366224 (accessed 16 September 2025)

67 United Nations Department of Economic and Social Affairs. The 17 goals | Sustainable Development. https://sdgs.un.org/goals (accessed 24 October 2025)

68 World Health Organization. Interruption of transmission and elimination of leprosy disease. 2023. https://www.who.int/publications/i/item/9789290210467 (accessed 24 October 2025)

69 World Health Organization African Region. WHO Regional Office for Africa / Press release | African health experts commit to accelerate efforts to eliminate Neglected Tropical Diseases. 2025. https://who-africa.africa-newsroom.com/press/african-health-experts-commit-to-accelerate-efforts-to-eliminate-neglected-tropical-diseases?lang=en (accessed 26 October 2025)

70 World Health Organization. Skin NTDs: prioritizing integrated approaches to reduce suffering, psychosocial impact and stigmatization. 2020. https://www.who.int/news/item/29-10-2020-skin-ntds-prioritizing-integrated-approaches-to-reduce-suffering-psychosocial-impact-and-stigmatization (accessed 25 November 2025)

71 Yeshanh WE. Scabies and podoconiosis: Entry points to demonstrate the feasibility of community based interventions for skin neglected tropical diseases. 2022. https://dare.uva.nl/search?identifier=0a131e29-3f8b-4019-a505-ce01103c8cfe (accessed 21 October 2025)

72 The Tanzania Society for Dermatovenereology (TASOD). TASOD - About us. https://www.tasod.or.tz/about-us (accessed 16 September 2025)

73 Bickers DR, Lim HW, Margolis D, et al. The burden of skin diseases: 2004. A joint project of the American Academy of Dermatology Association and the Society for Investigative Dermatology. J Am Acad Dermatol. 2006;55:490– 500. doi: 10.1016/j.jaad.2006.05.048

74 McCleskey PE, Gilson RT, DeVillez RL. Medical Student Core Curriculum in Dermatology Survey. J Am Acad Dermatol. 2009;61:30-35.e4. doi: 10.1016/J.JAAD.2008.10.066

75 Mwageni N, van Wijk R, Schoenmakers A, et al. The Reliability of Clinical Judgments in Dermatology: A Descriptive Review on Their Role as a Reference Standard for Technological Diagnostic Research. Under review.

76 Mwageni N, Schoenmakers A, van Wijk R, et al. From Rash to Diagnosis: First Insights on Capacity Strengthening of Primary Healthcare Workers by Formal and Informal Learning in Tanzanian “Skin Camps”. Publication expected after acceptance in Community Skin Health J. 2025.

77 Mieras L. PEP4LEP research and its contribution to capacity building. Lepr Rev. 2022;93:180–3. doi: 10.47276/LR.93.3.180

78 Urgesa K, de Bruijne ND, Bobosha K, et al. Prolonged delays in leprosy case detection in a leprosy hot spot setting in Eastern Ethiopia. PLoS Negl Trop Dis. 2022;16:e0010695. doi: 10.1371/JOURNAL.PNTD.0010695

79 Mwageni N, Kamara D, Kisonga R, et al. Leprosy epidemiological trends and diagnosis delay in three districts of Tanzania: A baseline study. Lepr Rev. 2022;93:209–23. doi: 10.47276/LR.93.3.209

80 Figueroa J. Prevalence of skin diseases in school children in rural and urban communities in the Illubabor province, south-western Ethiopia: a preliminary survey. J Eur Acad Dermatol Venereol. 1997;9:142–8. doi: 10.1016/S0926-9959(97)00105-0

81 Hogewoning A, Amoah A, Bavinck JNB, et al. Skin diseases among schoolchildren in Ghana, Gabon, and Rwanda. Int J Dermatol. 2013;52:589–600. doi: 10.1111/j.1365-4632.2012.05822.x

82 Tran H, Chen K, Lim AC, et al. Assessing diagnostic skill in dermatology: a comparison between general practitioners and dermatologists. Australas J Dermatol. 2005;46:230–4. doi: 10.1111/j.1440-0960.2005.00189.x

83 Federman DG, Concato J, Kirsner RS. Comparison of dermatologic diagnoses by primary care practitioners and dermatologists. A review of the literature. Arch Fam Med. 1999;8:170–2. doi: 10.1001/archfami.8.2.170

84 Peters DH, Tran NT, Adam T. Implementation Research in Health: A Practical Guide. 2013. https://iris.who.int/server/api/core/bitstreams/08869e5c-f0e0-49d2-903e-2f85a6336d99/content (accessed 25 November 2025)

85 PEP4LEP 2.0 consortium. PEP4LEP (2.0) – Raw Data Depository. 2025. https://www.leprosy-information.org/resource/pep4lep-20-raw-data-depository (accessed 26 November 2025)

